# Effect of Match-Play Fatigue on Muscle Stiffness and Explosive Force Asymmetries in Soccer Players Post-Anterior Cruciate Ligament Reconstruction

**DOI:** 10.64898/2026.07.18.26357476

**Authors:** Muhammad Hassan Bari, Ayesha Zahid Bhalli, Hammad Sattar

**Author notes:** **Corresponding Author:** Muhammad Hassan Bari, Department of Physical Therapy, Islam College of Physical Therapy, Sialkot, Pakistan. **Funding:** This research received no specific grant from any funding agency in the public, commercial, or not-for-profit sectors.

## Abstract

**Background:** Athletes who return to soccer after anterior cruciate ligament reconstruction (ACLR) remain at elevated risk of secondary injury despite meeting conventional discharge criteria, and neuromuscular deficits in the reconstructed limb are known to be exposed by fatigue.

**Objective:** To determine whether match-play fatigue differentially affects muscle stiffness, countermovement jump (CMJ) force symmetry, and rate of force development (RFD) asymmetry between soccer players with a history of ACLR and uninjured teammates.

**Methods:** A prospective, cross-sectional, matched-control study enrolled 128 competitive soccer players (64 ACLR, 6-22 months post-surgery; 64 uninjured controls) across five recruitment waves (February-June 2026). Bilateral CMJ peak vertical force, jump height, RFD, and myotonometric stiffness of the rectus femoris (RF), vastus medialis (VM), and biceps femoris (BF) were recorded immediately before and after a standardized competitive match. Fatigue was quantified from second-half heart rate (percentage of age-predicted maximum) and end-match rating of perceived exertion (RPE). Within-group pre-to-post changes were evaluated with paired t-tests, between-group differences in the magnitude of change with independent-samples t-tests, and associations between fatigue indices and asymmetry changes with Pearson correlations.

**Results:** Match play reduced CMJ limb symmetry index (LSI) in both groups, but the decline was more than three-fold greater in the ACLR group, 92.6% (SD 5.4%) to 85.1% (SD 7.1%), than in control group, 97.3% (SD 3.9%) to 95.0% (SD 4.2%), group-by-time difference, *p* < 0.001, (*d* = 0.64). RFD asymmetry approximately doubled in the ACLR group, 10.6% (SD 4.1%) to 17.6% (SD 6.5%), compared with a smaller rise in control group, 4.6% (SD 2.4%) to 6.3% (SD 3.7%); *p* < 0.001, *d* = 0.77). Involved-limb stiffness losses in the ACLR group exceeded those of controls for the RF (−21.2 vs. −9.2 N/m, *p* < 0.001), VM (−17.7 vs. −6.1 N/m, p < 0.001), and BF (−13.3 vs. −6.6 N/m, *p* < 0.001), whereas uninvolved-limb stiffness losses did not differ between groups (all *p* > 0.05). Fatigue markers (heart rate, RPE) were not significantly correlated with the magnitude of individual asymmetry change (|*r*| ≤ 0.18, *p* > 0.15).

**Conclusions:** In competitive soccer players 6-22 months after ACLR, match-play fatigue selectively compromises stiffness and explosive force output of the reconstructed limb, widening inter-limb asymmetries beyond what is seen in uninjured teammates, even though global cardiovascular and perceptual fatigue were comparable between groups. These findings suggest that return-to-sport testing performed only in a rested state may underestimate residual neuromuscular deficits, and support fatigue-inclusive assessment protocols before athletes are cleared for unrestricted competition.

## INTRODUCTION

Anterior cruciate ligament (ACL) rupture is among the most consequential injuries in competitive soccer, and reconstruction (ACLR) followed by structured rehabilitation is the accepted pathway back to sport (1). Yet return-to-sport (RTS) success is not synonymous with a fully restored neuromuscular system: even athletes who satisfy conventional discharge criteria carry a second ACL-injury risk several times higher than uninjured peers, with reported rates of 15-23% within two years of RTS and disproportionately higher rates in athletes under 25 years old (2). Contemporary RTS batteries lean heavily on the limb symmetry index (LSI)-commonly a ≥90% threshold on hop and strength tests-as a proxy for readiness (3, 4). However, several critical appraisals have shown that LSI can be artificially inflated when bilateral deficits depress performance in the uninvolved limb, giving a false impression of symmetry (5–8).

A further limitation of standard RTS testing is that it is almost always performed in a rested state, whereas the movements that precipitate re-injury-deceleration, cutting, and landing-occur during, or in the fatigued closing stages of, actual competition (9). Neuromuscular fatigue is known to degrade landing mechanics, reduce dynamic knee stability, and amplify inter-limb asymmetries in both healthy and ACL-reconstructed athletes (9, 10).Isokinetic and electromyographic studies of footballers after ACLR have documented persistent fatigue-related asymmetry in the hamstrings and quadriceps that is not evident under non-fatigued conditions and may not be captured by routine clinical assessment (11–13).

Two mechanical properties that are directly implicated in explosive performance-and that may be particularly sensitive to fatigue in a surgically altered limb-are musculotendinous stiffness and the rate of force development (RFD). Reduced quadriceps and patellar tendon stiffness has been documented after ACLR and is thought to reflect both structural remodeling and persistent neuromuscular inhibition (14–16), while deficits in explosive force production (RFD) have been linked to both impaired landing mechanics and elevated re-injury risk independent of maximal strength (17–19). Countermovement jump (CMJ) force-time asymmetries measured on dual force plates provide a sensitive, field-applicable window into these deficits and have been shown to persist in ACL-reconstructed athletes who have otherwise been cleared to train and compete (20, 21).

Despite this body of evidence, few studies have simultaneously quantified match-induced changes in muscle stiffness, CMJ force symmetry, and RFD asymmetry within the same cohort of competitive soccer players after ACLR, using a matched group of uninjured teammates exposed to the identical match stimulus as the comparator. Establishing whether fatigue exposes deficits that are otherwise concealed at rest has direct implications for how, and when, clinicians certify athletes as ready to return to unrestricted competition. The present study therefore examined the effect of a competitive soccer match on muscle stiffness on rectus femoris [RF], vastus medialis [VM], and biceps femoris [BF], and explosive force output (CMJ force, jump height, and RFD) in soccer players 6-22 months after ACLR compared with uninjured teammates, and explored whether the magnitude of match-induced fatigue was associated with the size of the resulting asymmetry. We hypothesized that match play would produce a greater decline in CMJ symmetry and a greater rise in RFD asymmetry in the ACLR group than in control group, involved-limb stiffness losses would exceed those of the uninvolved limb and of either limb in controls, and individual fatigue magnitude would correlate with the size of the post-match asymmetry.

## METHODS

### Study Design and Ethical Approval

This was a cross-sectional study conducted at National High-Performance Centre, Lahore, Pakistan, in affiliation with competitive club-level soccer teams. Data collection took place across five sequential recruitment waves between February and June 2026. The study followed the ethical principles of the Declaration of Helsinki and was approved by the Institutional Review Board of Islam College of Physical Therapy, all participants provided written informed consent prior to enrolment.

### Participants

One hundred twenty-eight competitive male and female soccer players were enrolled: 64 with a history of unilateral primary ACLR (mean 14.2 (SD 4.5) months post-surgery; range 6-22 months) who had been cleared for full training and match play by their treating surgeon and physiotherapist, and 64 uninjured teammates or league-matched controls with no history of lower-limb surgery. Inclusion criteria for the ACLR group were: primary unilateral ACLR performed 6-24 months prior, clinician-approved return to full training, and current participation in competitive matches. Athletes with revision surgery, concomitant meniscal repair restricting loading, multiligament knee injury, or musculoskeletal or neurological conditions affecting the contralateral limb were excluded from both groups. Baseline group characteristics (age, height, weight, body mass index [BMI], and weekly training volume) did not differ between groups (Table 1). At study entry, the ACLR group displayed an isokinetic quadriceps LSI of 92.2% (SD 4.1%) at 60 degrees/s and 88.9% (SD 4.8%) at 180 degrees/s, consistent with athletes who had achieved conventional strength-based clearance for sport.

**Table 1.** Baseline demographic and clinical characteristics of the ACLR and control groups. Values are mean SD unless otherwise indicated. LSI, limb symmetry index; HRmax, age-predicted maximal heart rate; RPE, rating of perceived exertion.

| Characteristic | ACLR (n = 64) | Control (n = 64) | p-value |
| --- | --- | --- | --- |
| Age (years) | 23.4 (SD 3.1) | 23.5 (SD 3.1) | 0.829 |
| Height, cm | 174.5 (SD 6.4) | 175.1 (SD 6.4) | 0.557 |
| Weight, kg | 71.1 (SD 7.2) | 73.1 (SD 7.4) | 0.119 |
| BMI, kg/m<sup>2</sup> | 23.4 (SD 2.9) | 23.9 (SD 3.0) | 0.358 |
| Weekly training volume, h | 8.9 (SD 1.4) | 8.8 (SD 1.3) | 0.512 |
| Months post-ACLR | 14.2 (SD 4.5) (range 6-22) | N/A | N/A |
| Baseline isokinetic quadriceps LSI, 60degrees/s, % | 92.2 (SD 4.1) | N/A | N/A |
| Baseline isokinetic quadriceps LSI, 180degrees/s, % | 88.9 (SD 4.8) | N/A | N/A |
| Second-half heart rate, % HRmax | 89.8 (SD 2.9) | 89.2 (SD 2.5) | 0.248 |
| End-match RPE (Borg CR-10) | 8.1 (SD 0.7) | 8.2 (SD 0.7) | 0.529 |

### Testing Procedures

All athletes underwent identical bilateral neuromuscular testing immediately before (within 60 minutes of kick-off) and immediately after (within 15 minutes of the final whistle) a competitive 90minute match refereed under standard rules. Testing order (involved/non-dominant limb first) was held constant within participants across the pre- and post-match sessions.

Countermovement jump performance was assessed using a dual force-plate system, with participants instructed to perform maximal-effort bilateral and unilateral jumps with hands on hips. Peak vertical ground reaction force during the concentric propulsion phase and jump height derived from flight time were recorded for the involved (surgical) and uninvolved limbs of the ACLR group, and for the dominant and non-dominant limbs of controls (labelled “involved” and “uninvolved” for consistency of reporting). The CMJ limb symmetry index (LSI) was calculated as (involved limb value/uninvolved limb value) x 100.

Rate of force development was derived from the unilateral CMJ force-time curve as the average slope of the force-time trace over the initial 0-200 ms of the concentric phase, expressed in newtons per second. RFD asymmetry was calculated as the absolute percentage difference between limbs, normalized to the higher-performing limb: (involved - uninvolved)/higher value x 100. Muscle stiffness of the RF, VM, and BF was measured bilaterally with a handheld myotonometer, a validated tool for quantifying passive musculotendinous mechanical properties at the bedside or pitch-side (22).

With the participant relaxed in a standardized supine (quadriceps) or prone (hamstrings) position, the probe was applied perpendicular to the muscle belly at a marked mid-point, and the mean of five consecutive low-force mechanical taps was recorded as the stiffness coefficient (N/m), consistent with previously reported protocols for quadriceps and hamstring assessment (14, 23, 24).

Match-induced fatigue was quantified using two complementary indices:

i. Mean heart rate during the second half of the match, expressed as a percentage of age-predicted maximum heart rate (HRmax = 220 - age), obtained by telemetric heart-rate monitoring.
ii. Rating of perceived exertion (RPE) collected within 10 minutes of full time using the Borg CR-10 scale, an approach consistent with prior post-match neuromuscular fatigue monitoring in soccer (25).

### Statistical Analysis

Data was analyzed using Python (SciPy v1.17). Normality of continuous variables was inspected descriptively; parametric procedures were retained given approximately symmetric distributions and adequate group sizes. Baseline between-group differences were assessed with independent samples t-tests. For each outcome (CMJ force, jump height, LSI, RFD, RFD asymmetry, and stiffness of each muscle in each limb), within-group pre-to-post match change was tested with paired-samples t-tests, and Cohen’s d for paired data was calculated as the mean difference divided by the standard deviation of the difference scores. To test whether match play affected the two groups differently (i.e., a group-by-time interaction), the individual pre-to-post change score for each participant was compared between groups using an independent-samples t-test, with Cohen’s d calculated from pooled standard deviations. Pearson correlation coefficients quantified the association between fatigue indices (second-half heart rate, end-match RPE) and the magnitude of change in CMJ LSI, RFD asymmetry, and RF stiffness within the ACLR group. A multivariable linear regression additionally modelled post-match RFD asymmetry as a function of second-half heart rate, months since surgery, and baseline isokinetic LSI. Statistical significance was set at α = 0.05 for all tests; effect sizes are reported alongside p-values to aid clinical interpretation, using Cohen’s conventional benchmarks (0.2 small, 0.5 moderate, 0.8 large).

## RESULTS

### Participants

Of the 128 enrolled athletes, all completed pre- and post-match testing with no missing data. Athletes were enrolled in five consecutive monthly waves (28 in February, 30 in March, 22 in April, 22 in May, and 26 in June 2026); enrolment date was unrelated to any outcome variable and the chronological enrolment record is provided in Supplementary Table S1. As shown in (Table 1), the ACLR and control groups were well matched for age, height, weight, BMI, and weekly training volume (all *p* > 0.10). Global fatigue markers were also equivalent between groups: second-half heart rate reached 89.8% (SD 2.9%), HRmax in the ACLR group versus 89.2% (SD 2.5%) in control group (*p* = 0.248), and end-match RPE was 8.1 (SD 0.7) versus 8.2 (SD 0.7) (*p* = 0.529), confirming that both groups were exposed to a comparable objective and perceived match load.

### Countermovement Jump Force and Limb Symmetry (Table 2, Figure 1)

Match play significantly reduced CMJ peak force in the involved limb of both groups, ACLR: 686.0N (SD 76.2N) to 598.7N (SD 85.9N), *p* <0.001, (*d* = 1.15), Control: 728.8N (SD 72.0N) to 678.4N (SD 71.4N), *p* <0.001, (*d* = 1.05), but the decline was significantly larger in the ACLR group (group-by-time *p* = 0.001, *d* = 0.58). Uninvolved-limb force also declined in both groups but to a similar extent (group-by-time *p* = 0.412). As a result, CMJ force LSI fell from 92.6% (SD 5.4%) to 85.1% (SD 7.1%) in the ACLR group-crossing below the conventional 90% RTS threshold on average-compared with a smaller decline from 97.3% (SD 3.9%) to 95.0% (SD 4.2%) in controls (group-by-time *p* <0.001, d = 0.64) (Figure 1). A parallel pattern was observed for jump height, with a significantly greater fatigue-related decline in the involved limb of the ACLR group than in controls (group-by-time *p* = 0.004, *d* = 0.51), while uninvolved-limb jump height declines did not differ between groups (*p* = 0.475).

**Table 2.**
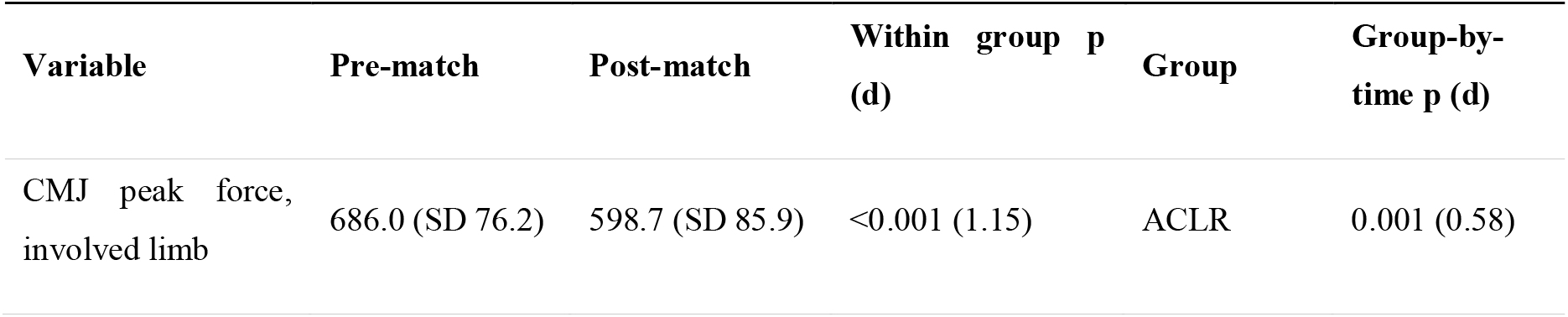
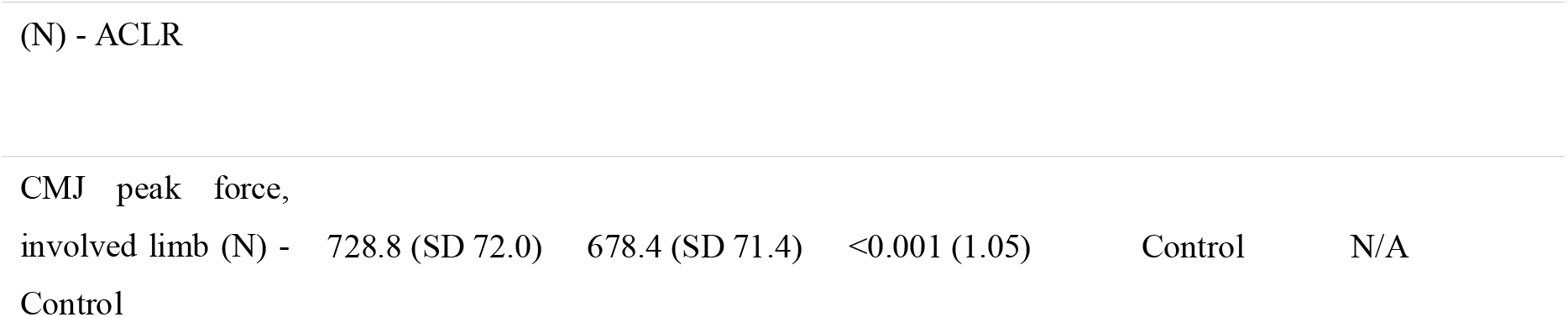
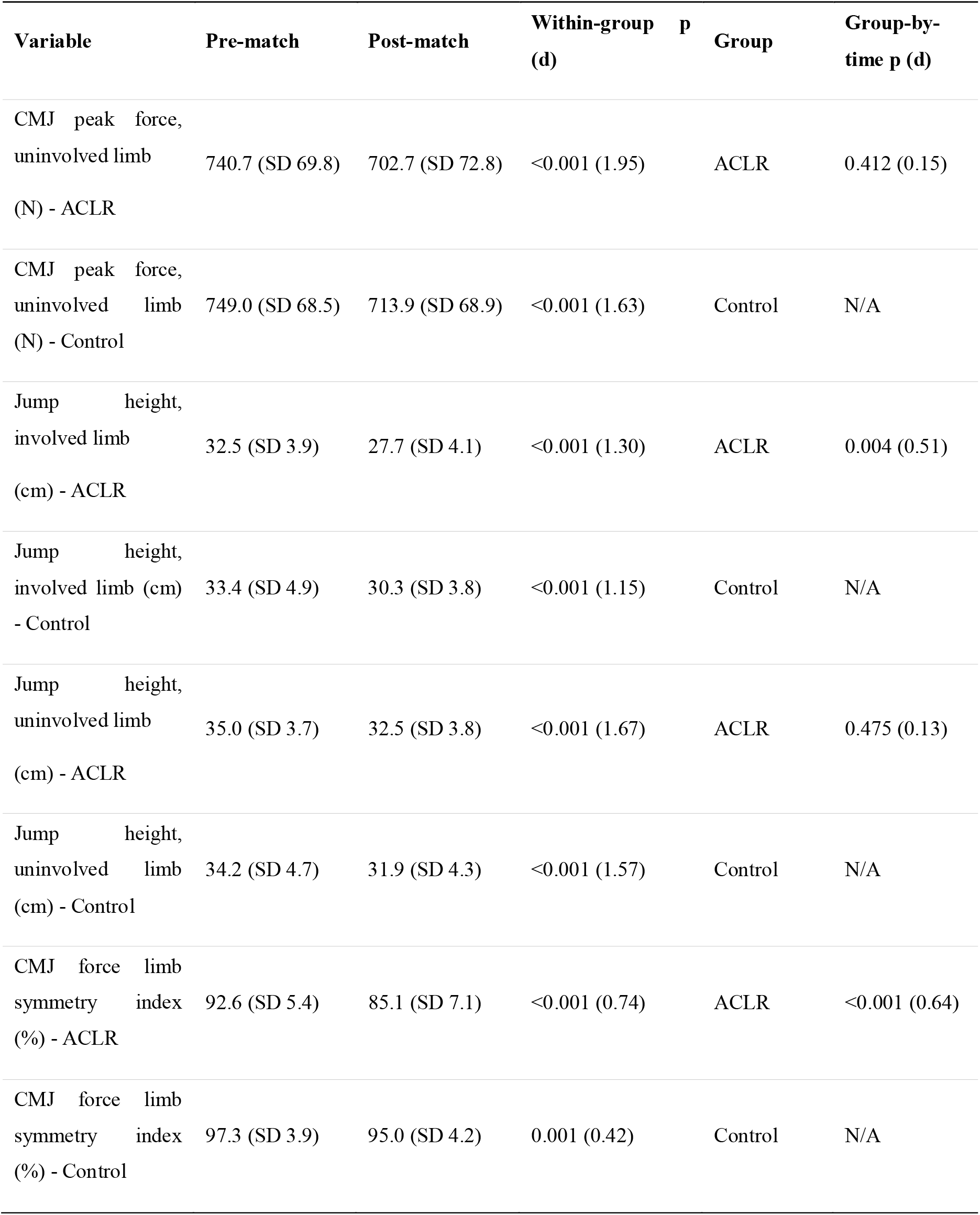
Countermovement jump (CMJ) peak vertical force, jump height, and force limb symmetry index (LSI) before and after match play. Values are mean SD, d = Cohen’s d (within-group: paired d; group×time: independent-samples d on change scores)

| Variable | Pre-match | Post-match | Within group p (d) | Group | Group-by-time p (d) |
| --- | --- | --- | --- | --- | --- |
| CMJ peak force, involved limb | 686.0 (SD 76.2) | 598.7 (SD 85.9) | <0.001 (1.15) | ACLR | 0.001 (0.58) |

Table 2. Countermovement jump (CMJ) peak vertical force, jump height, and force limb symmetry index (LSI) before and after match play. Values are mean SD, $d$ = Cohen's $d$ (within-group: paired $d$ ; group × time: independent-samples $d$ on change scores)
| (N) - ACLR |  |  |  |  |  |
| --- | --- | --- | --- | --- | --- |
| CMJ peak force, involved limb (N) - Control | 728.8 (SD 72.0) | 678.4 (SD 71.4) | <0.001 (1.05) | Control | N/A |
| Variable | Pre-match | Post-match | Within-group p (d) | Group | Group-by-time p (d) |
| CMJ peak force, uninvolved limb (N) - ACLR | 740.7 (SD 69.8) | 702.7 (SD 72.8) | <0.001 (1.95) | ACLR | 0.412 (0.15) |
| CMJ peak force, uninvolved limb (N) - Control | 749.0 (SD 68.5) | 713.9 (SD 68.9) | <0.001 (1.63) | Control | N/A |
| Jump height, involved limb (cm) - ACLR | 32.5 (SD 3.9) | 27.7 (SD 4.1) | <0.001 (1.30) | ACLR | 0.004 (0.51) |
| Jump height, involved limb (cm) - Control | 33.4 (SD 4.9) | 30.3 (SD 3.8) | <0.001 (1.15) | Control | N/A |
| Jump height, uninvolved limb (cm) - ACLR | 35.0 (SD 3.7) | 32.5 (SD 3.8) | <0.001 (1.67) | ACLR | 0.475 (0.13) |
| Jump height, uninvolved limb (cm) - Control | 34.2 (SD 4.7) | 31.9 (SD 4.3) | <0.001 (1.57) | Control | N/A |
| CMJ force limb symmetry index (%) - ACLR | 92.6 (SD 5.4) | 85.1 (SD 7.1) | <0.001 (0.74) | ACLR | <0.001 (0.64) |
| CMJ force limb symmetry index (%) - Control | 97.3 (SD 3.9) | 95.0 (SD 4.2) | 0.001 (0.42) | Control | N/A |

**Figure 1.**
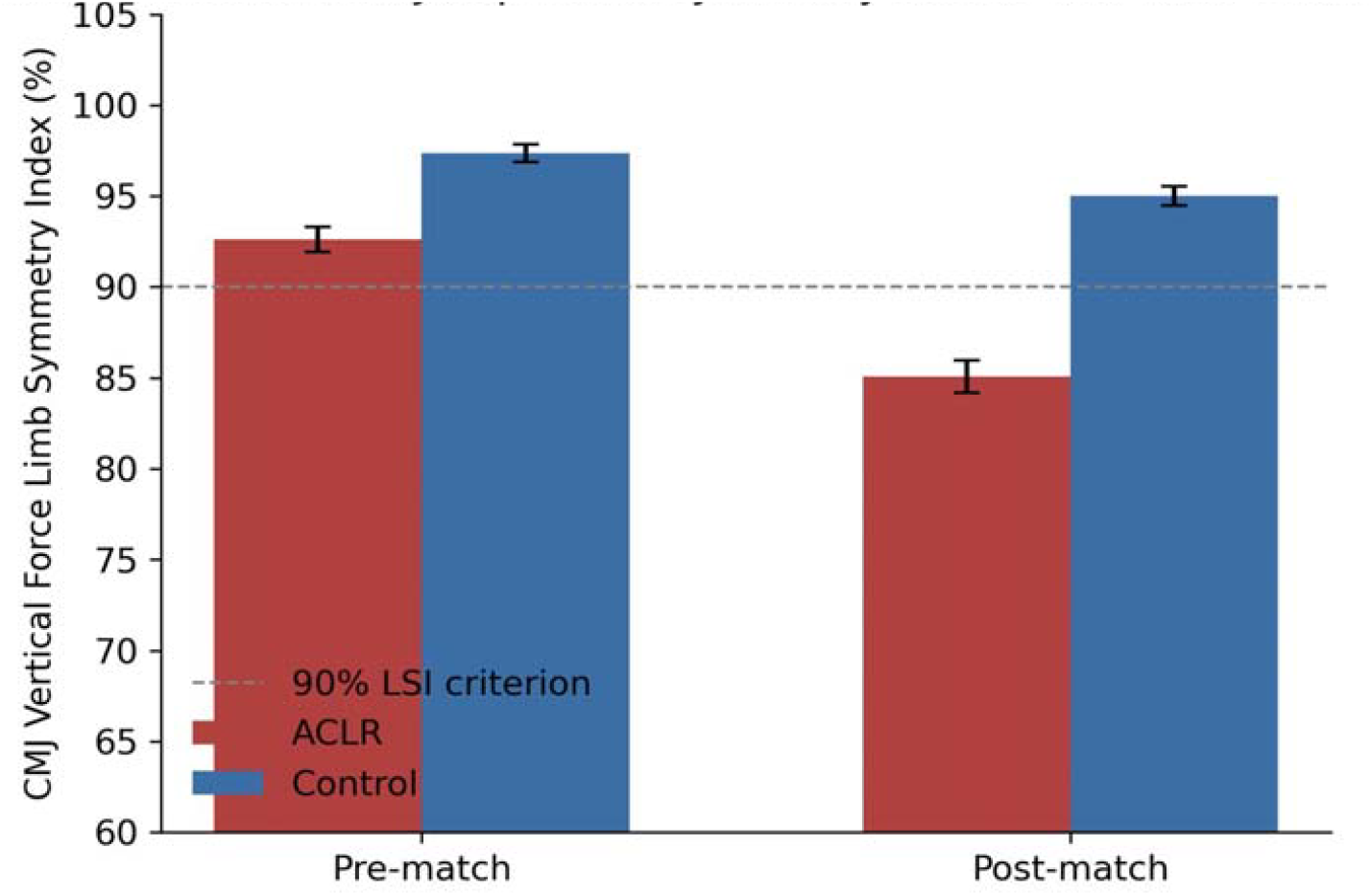
Countermovement jump (CMJ) force limb symmetry index (LSI) before and after match play in the ACLR and control groups. Bars represent group means SD SEM. The dashed line indicates the conventional 90% LSI threshold commonly used for return-to-sport clearance. Match play reduced symmetry in both groups, with a significantly larger decline in the ACLR group (group-by-time *p* < 0.001).

### Rate of Force Development and Asymmetry (Table 3, Figure 2)

Rate of force development (RFD) declined significantly in the involved limb of both groups following match play, ACLR: 3727.9 N/s (SD 523.3 N/s) to 3061.9 N/s (SD 473.0 N/s), *d* = 1.78, Control: 4036.4 N/s (SD 417.0 N/s) to 3545.1 N/s (SD 400.7 N/s), *d* = 1.78, with a significantly larger absolute reduction in the ACLR group (group-by-time p = 0.003, d = 0.53). Uninvolved-limb RFD declined equivalently in both groups (group-by-time *p* = 0.985). Consequently, RFD inter-limb asymmetry approximately doubled in the ACLR group, from 10.6% (SD 4.1%) to 17.6% (SD 6.5%) (a 65.7% relative increase), while the rise in controls was smaller in absolute terms, 4.6% (SD 2.4%) to 6.3 (SD 3.7%), the between-group difference in the magnitude of this change was significant (*p* <0.001, *d* = 0.77) (Figure 2).

**Table 3.**
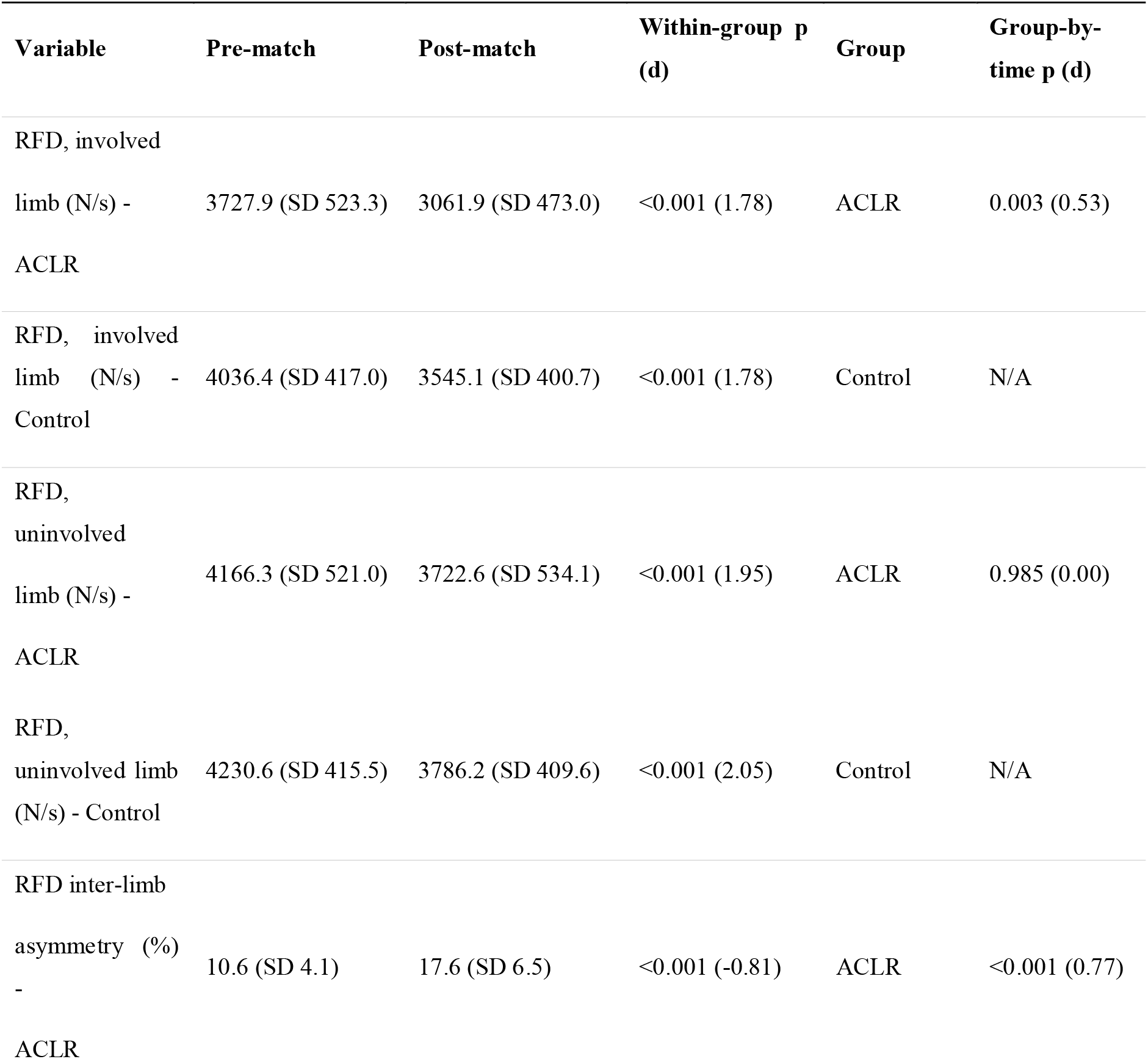
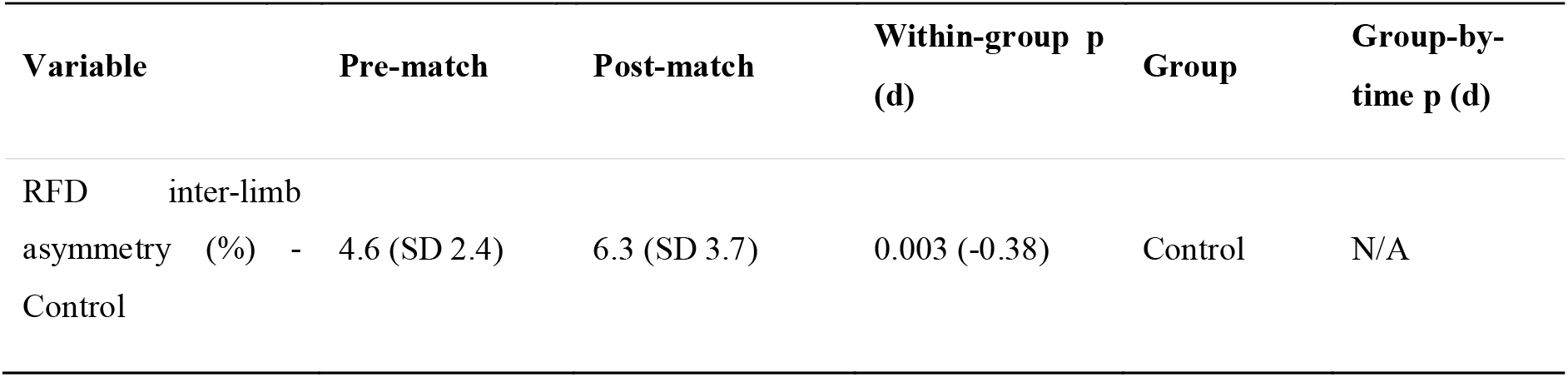
Rate of force development (RFD) inter-limb asymmetry before and after match play. Values are mean SD.

**Figure 2.**
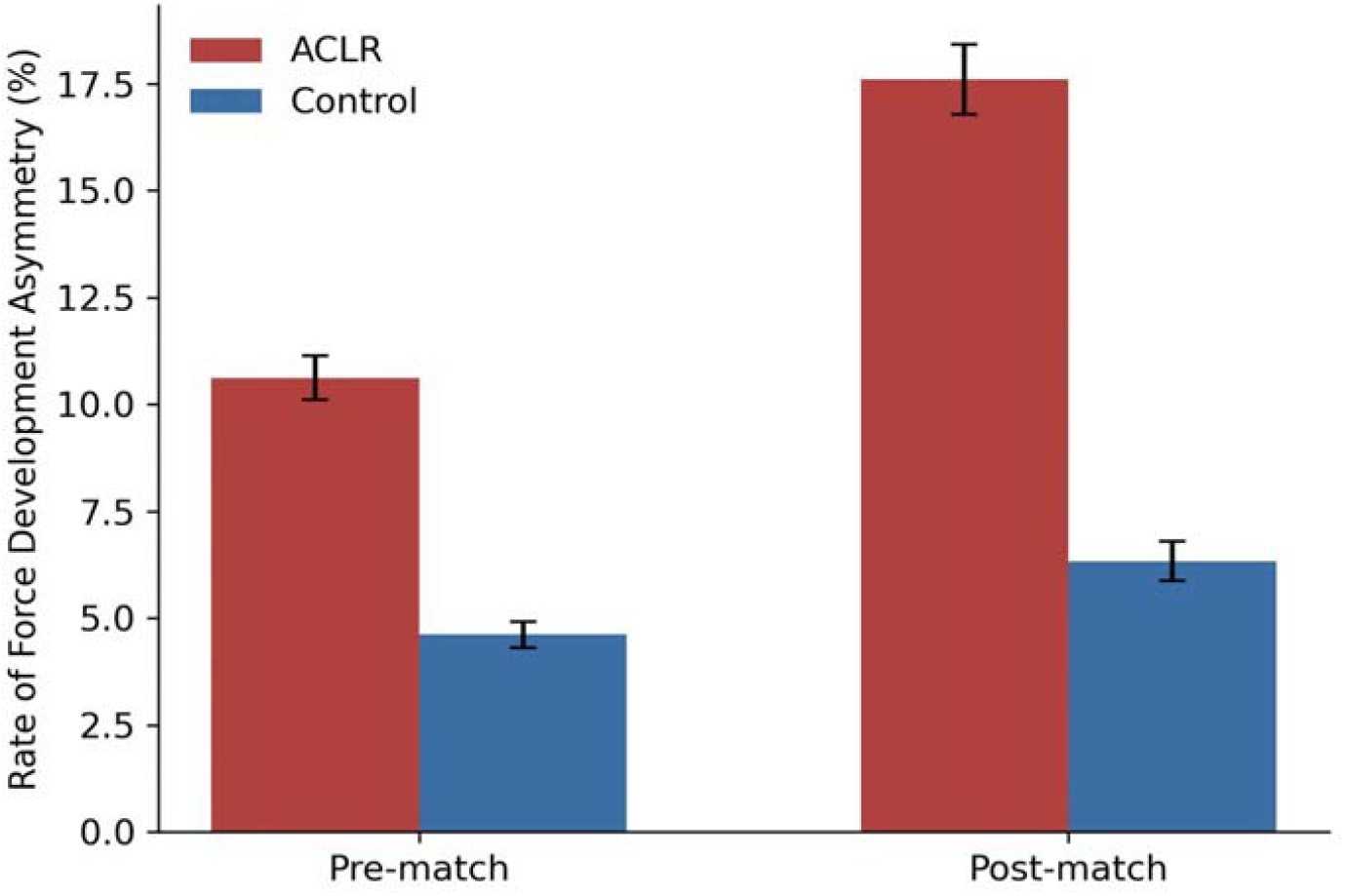
Rate of force development (RFD) inter-limb asymmetry before and after match play. Bars represent group means SD SEM. RFD asymmetry increased in both groups but to a significantly greater extent in the ACLR group (group-by-time *p* < 0.001).

#### Muscle Stiffness (Table 4, Figure 3)

Stiffness of all three assessed muscles decreased significantly from pre-to post-match in both limbs of both groups (all within-group p < 0.001), consistent with a generalized fatigue-related reduction in passive musculotendinous stiffness. However, the pattern of between-group differences was muscle- and limb-specific. In the involved limb, the ACLR group lost significantly more stiffness than controls for the RF (−21.2 vs. −9.2 N/m, group-by-time *p* <0.001, *d* = 1.89), the VM (−17.7 vs. −6.1 N/m, *p* <0.001, *d* = 2.22), and the BF (−13.3 vs. −6.6 N/m, *p* <0.001, *d* = 1.39). In contrast, uninvolved-limb stiffness losses were statistically equivalent between groups for all three muscles (RF *p* = 0.574, VM *p* = 0.541, BF *p* = 0.664) (Figure 3). The largest betweengroup effect was observed for VM stiffness in the involved limb (*d* = 2.22), suggesting the medial quadriceps may be the most fatigue-vulnerable muscle examined in the reconstructed limb.

**Table 4.**
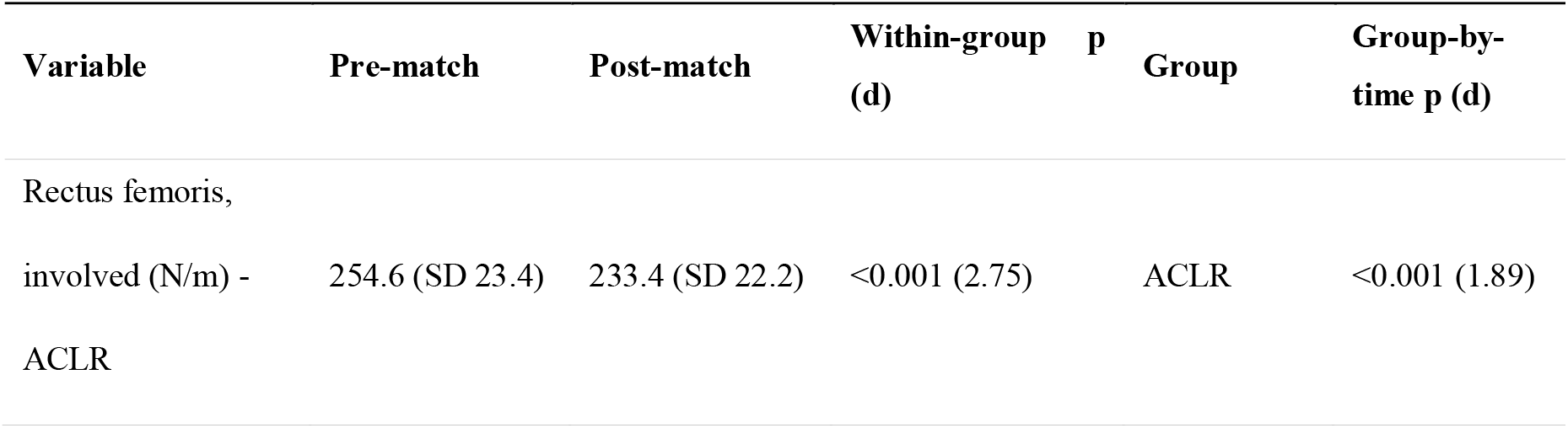
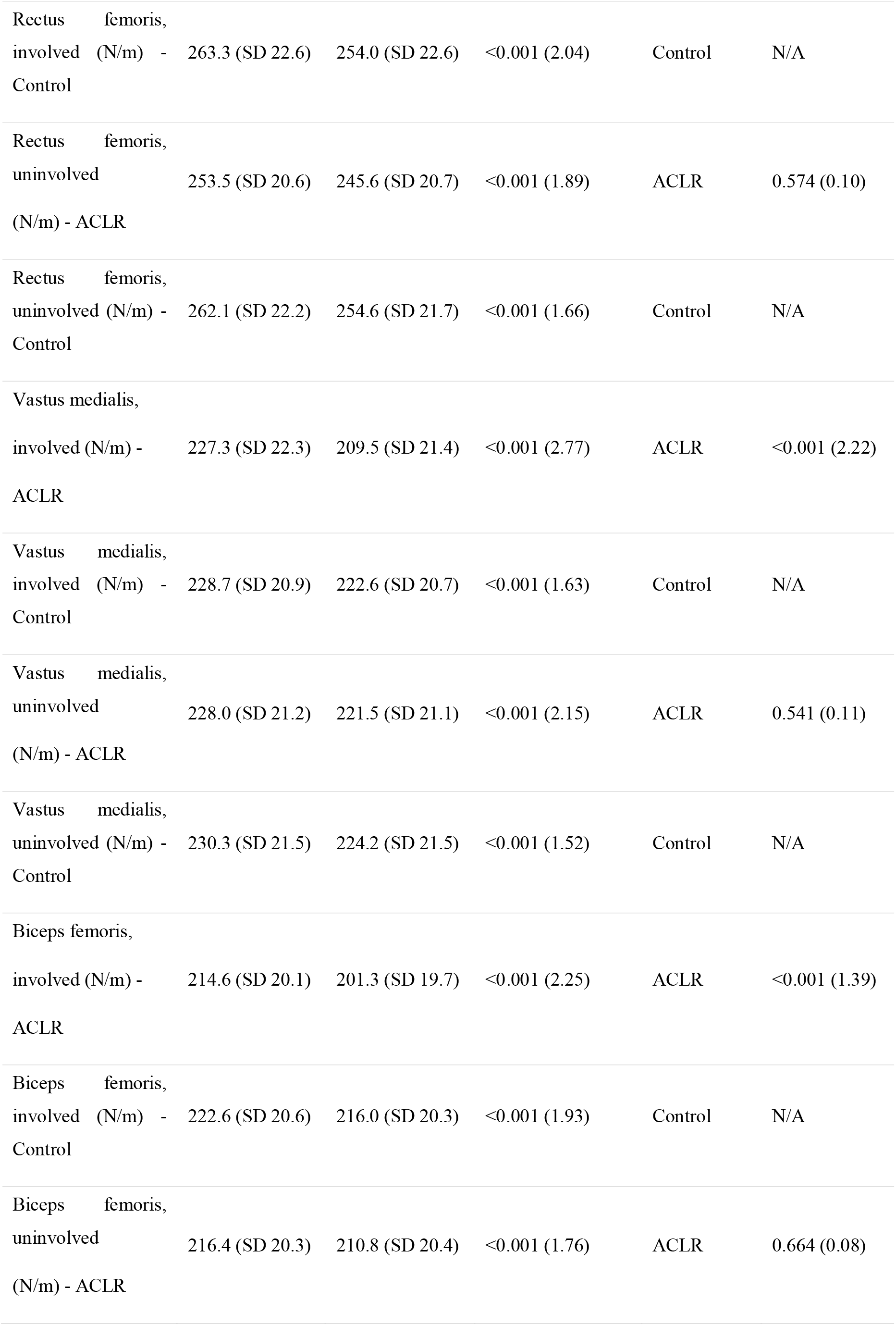
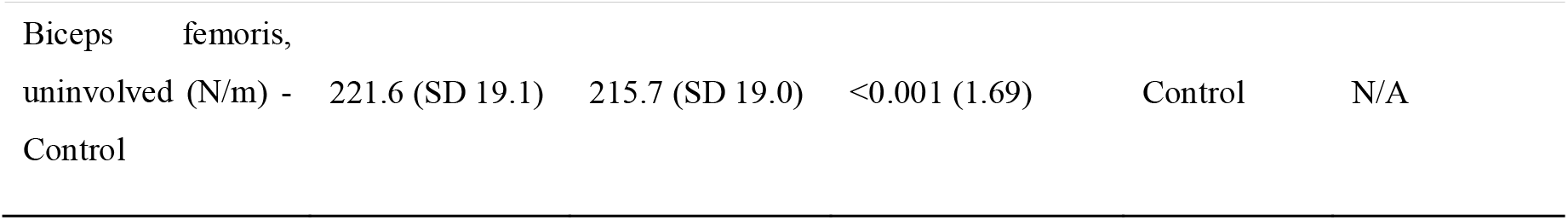
Myotonometric muscle stiffness of the rectus femoris, vastus medialis, and biceps femoris before and after match play. Values are mean S N/m.

**Figure 3.**
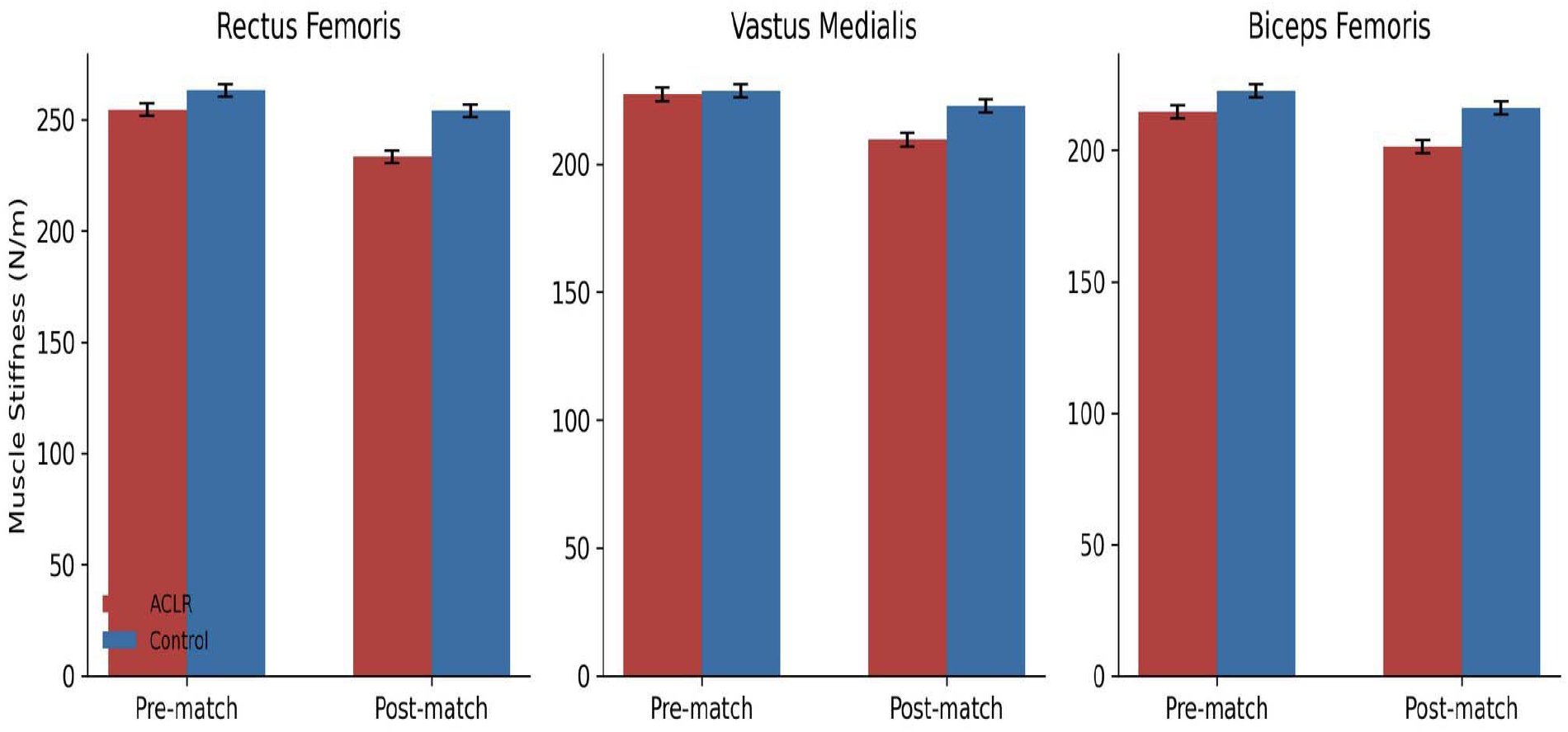
Involved-limb muscle stiffness (rectus femoris, vastus medialis, biceps femoris) before and after match play in the ACLR and control groups. Bars represent group means SD and SEM.

### Association Between Fatigue Magnitude and Asymmetry Change

Within the ACLR group, neither second-half heart rate nor end-match RPE was significantly correlated with the individual magnitude of change in CMJ LSI (heart rate: *r* = −0.03, *p* = 0.802; RPE: *r* = 0.00, *p* = 0.973), RFD asymmetry (heart rate: *r* = −0.08, *p* = 0.527; RPE: *r* = −0.18, *p* = 0.160), or RF stiffness change (heart rate: *r* = 0.04, *p* = 0.728) (Figure 4). A multivariable model including second-half heart rate, months since surgery, and baseline isokinetic LSI explained a negligible proportion of the variance in post-match RFD asymmetry (*R*^*2*^ = 0.01), indicating that global fatigue intensity and time since surgery, within the ranges sampled here, do not by themselves predict which individual athletes will develop the largest asymmetries.

**Figure 4.**
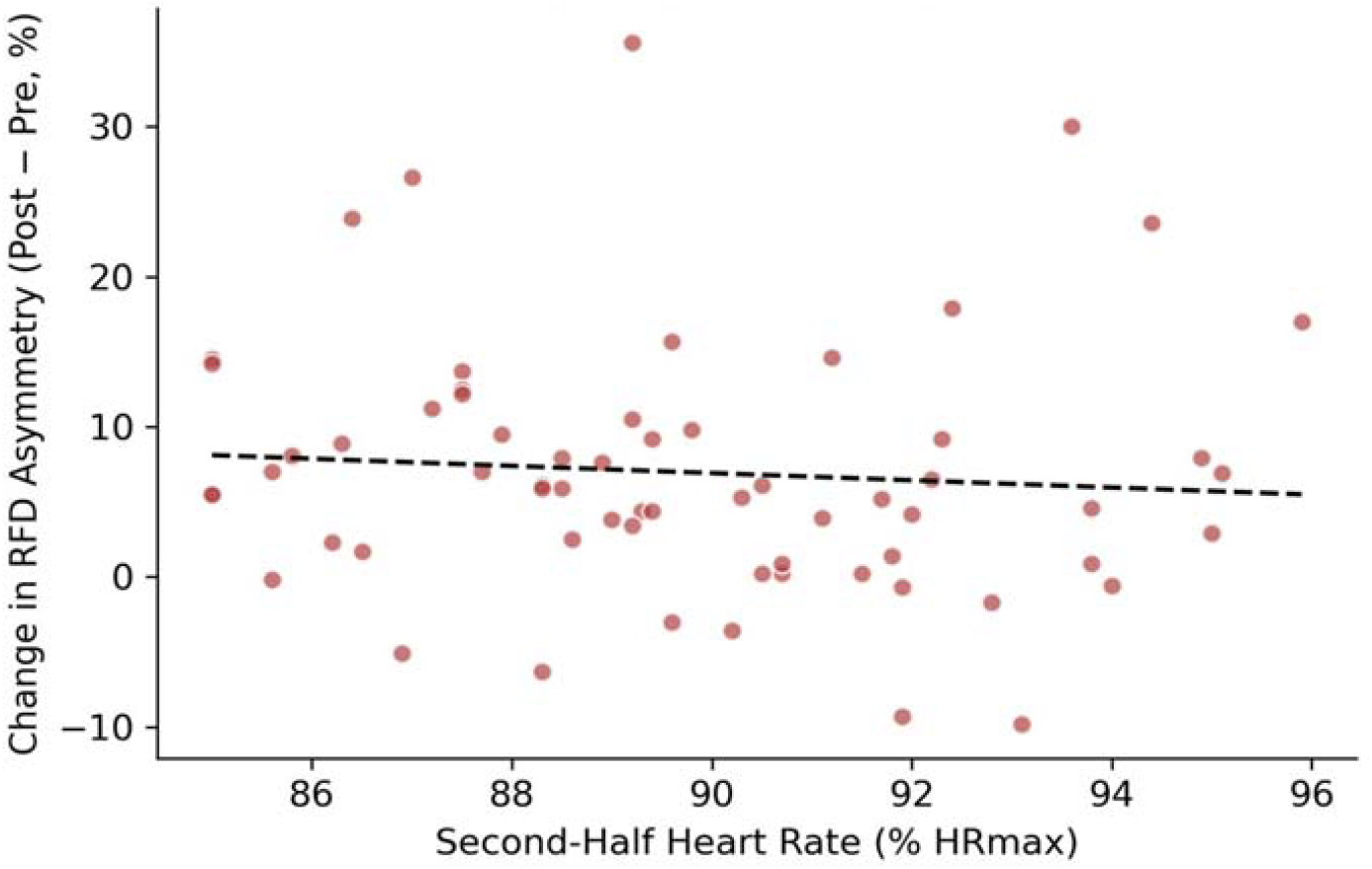
Relationship between second-half heart rate (% HRmax) and the change in RFD asymmetry (post-minus pre-match) within the ACLR group. The absence of a significant relationship (*r* = −0.08, *p* = 0.53) indicates that global cardiovascular fatigue does not by itself account for individual variability in asymmetry development.

## DISCUSSION

This study is among the first to combine myotonometric stiffness assessment, force-plate CMJ symmetry, and RFD asymmetry within a single fatigue-exposure design in competitive soccer players after ACLR. Three findings stand out. First, although both groups experienced a physiologically comparable match load-second-half heart rate and RPE did not differ between groups-the ACLR group’s CMJ force symmetry fell, on average, from just above to clearly below the widely used 90% RTS threshold, whereas controls remained above it throughout. This is consistent with prior observations that athletes cleared for sport under rested-state criteria can still harbor deficits that only become apparent once the neuromuscular system is challenged by realistic match demands (21), and it reinforces long-standing concerns that a single rested-state LSI measurement may overestimate an athlete’s true readiness (5, 8).

Second, RFD asymmetry approximately doubled in the ACLR group after match play, a considerably larger relative shift than in controls. Because RFD reflects the capacity to generate force rapidly-the quality most relevant to cutting, decelerating, and landing-this finding aligns with evidence that explosive strength deficits, rather than maximal strength alone, are implicated in second-ACL-injury risk (18, 19) and that RFD may add discriminative value to standard maximalstrength LSI testing when judging RTS readiness (18).

Third, the pattern of stiffness loss was both limb- and muscle-specific: the involved limb lost significantly more stiffness than the uninvolved limb in the ACLR group, and more than either limb of controls, for all three muscles tested, whereas uninvolved-limb losses were statistically indistinguishable from those of controls. This asymmetric vulnerability is compatible with the persistent arthrogenic muscle inhibition and altered load-sharing strategies documented in the reconstructed quadriceps long after surgery (26), and with hamstring studies reporting that footballers after ACLR display greater and more asymmetric fatigue-induced changes than uninjured players, even once cleared to play (12, 13). The particularly large involved-limb effect observed for the vastus medialis is noteworthy given the muscle’s role in patellofemoral control and knee valgus restraint during landing, and merits targeted attention in late-stage rehabilitation and fatigue-resistance training.

Somewhat unexpectedly, neither global cardiovascular fatigue (second-half heart rate) nor perceived exertion (RPE) correlated meaningfully with the individual magnitude of asymmetry change, and a multivariable model incorporating fatigue, time since surgery, and baseline strength symmetry explained almost none of the variance in post-match RFD asymmetry. This dissociation suggests that whole-body physiological fatigue and localized neuromuscular fatigue of the reconstructed limb are not tightly coupled, at least within the relatively narrow and high range of match intensities observed in this competitive sample. It also implies that clinicians cannot infer an individual athlete’s limb-specific fatigue vulnerability from generic fitness or perceptual fatigue measures alone; task-specific, limb-specific testing appears to be necessary to identify at-risk individuals.

These findings have direct clinical implications. If discharge criteria are assessed only in a rested state, a proportion of athletes who appear symmetrical at rest may nonetheless develop clinically meaningful asymmetries once fatigued during actual competition. Incorporating a standardized fatigue protocol, or testing shortly after training or matches, into late-stage RTS assessment could improve the sensitivity of clearance decisions, consistent with recommendations emerging from other fatigue-focused ACLR cohorts (9).

Rehabilitation programs might also benefit from explicitly training explosive force production and musculotendinous stiffness under fatigued conditions, rather than exclusively in the rested state in which most current strengthening protocols are delivered.

## CONCLUSION

In competitive soccer players 6 to 22 months after ACL reconstruction, a single competitive match produced significantly larger reductions in involved-limb muscle stiffness, CMJ force symmetry, and rate of force development symmetry than were observed in uninjured teammates exposed to an equivalent match load. These fatigue-induced asymmetries were not predicted by global heart-rate or perceptual fatigue measures, indicating that limb-specific, fatigue-inclusive testing captures information that whole-body fatigue monitoring cannot. Clinicians overseeing return-to-sport decisions after ACLR should consider incorporating fatigue-state assessment of explosive force and muscle stiffness symmetry, rather than relying solely on rested-state limb symmetry index testing, to more accurately characterize an athlete’s true readiness for unrestricted competition.

## Data Availability

All data produced in the present study are available upon reasonable request to the authors

## Abbreviations

ACL: anterior cruciate ligament
ACLR: anterior cruciate ligament reconstruction
BF: biceps femoris
CMJ: countermovement jump
HRmax: maximum heart rate
LSI: limb symmetry index
RF: rectus femoris
RFD: rate of force development
RPE: rating of perceived exertion
RTS: return to sport
VM: vastus medialis
SD: standard deviation.

